# Developing and Evaluating Pediatric Phecodes (Peds-Phecodes) for High-Throughput Phenotyping Using Electronic Health Records

**DOI:** 10.1101/2023.08.22.23294435

**Authors:** Monika E. Grabowska, Sara L. Van Driest, Jamie R. Robinson, Anna E. Patrick, Chris Guardo, Srushti Gangireddy, Henry Ong, QiPing Feng, Robert Carroll, Prince J. Kannankeril, Wei-Qi Wei

## Abstract

**Objective:** Pediatric patients have different diseases and outcomes than adults; however, existing phecodes do not capture the distinctive pediatric spectrum of disease. We aim to develop specialized pediatric phecodes (Peds-Phecodes) to enable efficient, large-scale phenotypic analyses of pediatric patients.

**Materials and Methods:** We adopted a hybrid data- and knowledge-driven approach leveraging electronic health records (EHRs) and genetic data from Vanderbilt University Medical Center to modify the most recent version of phecodes to better capture pediatric phenotypes. First, we compared the prevalence of patient diagnoses in pediatric and adult populations to identify disease phenotypes differentially affecting children and adults. We then used clinical domain knowledge to remove phecodes representing phenotypes unlikely to affect pediatric patients and create new phecodes for phenotypes relevant to the pediatric population. We further compared phenome-wide association study (PheWAS) outcomes replicating known pediatric genotype-phenotype associations between Peds-Phecodes and phecodes.

**Results:** The Peds-Phecodes aggregate 15,533 ICD-9-CM codes and 82,949 ICD-10-CM codes into 2,051 distinct phecodes. Peds-Phecodes replicated more known pediatric genotype-phenotype associations than phecodes (248 versus 192 out of 687 SNPs, p<0.001).

**Discussion:** We introduce Peds-Phecodes, a high-throughput EHR phenotyping tool tailored for use in pediatric populations. We successfully validated the Peds-Phecodes using genetic replication studies. Our findings also reveal the potential use of Peds-Phecodes in detecting novel genotype-phenotype associations for pediatric conditions. We expect that Peds-Phecodes will facilitate large-scale phenomic and genomic analyses in pediatric populations.

**Conclusion:** Peds-Phecodes capture higher-quality pediatric phenotypes and deliver superior PheWAS outcomes compared to phecodes.

## BACKGROUND AND SIGNIFICANCE

The pediatric spectrum of disease is distinct from its adult counterpart.[1] Children experience a variety of illnesses not commonly observed in adults. This includes congenital anomalies, genetic disorders with early mortality, and certain infectious diseases.[2,3] Conversely, adults experience many diseases unlikely to affect pediatric patients, such as Alzheimer’s disease, breast and prostate cancer, osteoarthritis, and many other conditions associated with aging.[4] Although pediatric data have rapidly accumulated in electronic health records (EHRs), pediatric patients have not been prioritized in developing high-throughput phenotyping tools such as phecodes, preventing researchers from performing focused large-scale analyses of pediatric data, including pediatric-specific phenome-wide association studies (PheWAS) and genome-wide association studies (GWAS), contributing to missed opportunities for scientific discovery.

The development of phecodes represents a key effort in EHR phenotyping. Phecodes aggregate relevant International Classification of Diseases (ICD-9-CM, ICD-10-CM, and ICD-10) codes into distinct phenotypes to better represent clinically meaningful diseases and traits (e.g., grouping ICD-9-CM codes 162.* representing lung cancer and ICD-9-CM codes V10.1* representing a history of lung cancer).[5,6] Phecodes are represented using numeric codes arranged in a three-level hierarchy, allowing phenotypes to be captured at various levels of granularity. Root phecodes, located at the top of the phecode hierarchy, provide the broadest phenotype definitions and are represented using whole numbers. These root phecodes can then branch into progressively more detailed sub-phecodes, indicated by decimal digits. In the current phecodes (version 1.2), up to two levels of additional phenotypic granularity (i.e., two decimal places) are available for each root phecode. Phecodes with a single digit following the decimal point are referred to as level 1 sub-phecodes; level 2 sub-phecodes have two digits after the decimal point. For example, root phecode 008 “Intestinal infection” branches into level 1 sub-phecode 008.5 “Bacterial enteritis”, which then branches into level 2 sub-phecode 008.51 “Intestinal E. coli” (Supplementary Figure 1).

We previously demonstrated that phecodes produced superior results in PheWAS compared to other coding systems, including ICD and the Clinical Classifications Software.[5] Since their introduction, phecodes have been globally used in multiple studies to both replicate known genotype-phenotype associations and discover new ones.[7–9] Phecodes have also been used beyond genetics to study long-term disease effects.[10] The latest version of phecodes can be found at: https://wei-lab.app.vumc.org/phecode-data/phecodes

While the existing phecodes are valuable in biomedical research, they need to be optimized for use in pediatric phenomic analyses. Phecodes were developed using population-based diagnoses predominantly from adult patients, which do not accurately reflect pediatric conditions. Modifying the existing phecodes to capture diseases primarily affecting pediatric patients with increased granularity and exclude age-related diseases uncommon in the pediatric population presents an opportunity to increase statistical power for identifying signals more efficiently and accurately.

In this study, we use current phecodes as a starting point to create Peds-Phecodes, specialized pediatric phecodes that more appropriately reflect the unique spectrum of pediatric disease, and evaluate their performance in large-scale PheWAS analyses using real-world data.

## MATERIALS AND METHODS

### Data source

We used de-identified EHRs from Vanderbilt University Medical Center’s (VUMC) Synthetic Derivative, a repository of rich, longitudinal clinical information encompassing data from >3.5 million patients, including data from >1 million pediatric individuals.[11] For our validation analyses, we also used data from VUMC’s DNA biobank, BioVU,[12] which links the genetic data of >100,000 individuals to their de-identified EHR data (>50,000 individuals with available pediatric EHR data).

### Pediatric phecode (Peds-Phecodes) development

We mapped ICD diagnoses captured while patients were <18 years (pediatric) and ≥18 years (adult) in VUMC’s EHR to current phecodes (N = 1,866) and compared the prevalence of each phecode in the pediatric and adult populations. We used the chi-square test to assess for significant differences in prevalence between the two groups if both proportions were over 5%. Otherwise, we used Fisher’s exact test to assess the difference. We used a Bonferroni-corrected significance threshold to adjust for multiple testing. Additionally, we flagged phecodes with low pediatric patient representation (N<50 pediatric patients) as phecodes for potential removal, as 50 cases has been suggested to be the minimum sample size required to detect an association in PheWAS analyses of binary traits.[13]

We adopted a hybrid data- and knowledge-driven approach to develop the pediatric phecodes (Peds-Phecodes) (Figure 1). Because of the three-tiered hierarchical structure of the phecodes (root phecodes, level 1 sub-phecodes, and level 2 sub-phecodes), an important component of this work involved ensuring that all modified phecodes continued to belong to the correct phecode “tree”.

**Figure 1.**
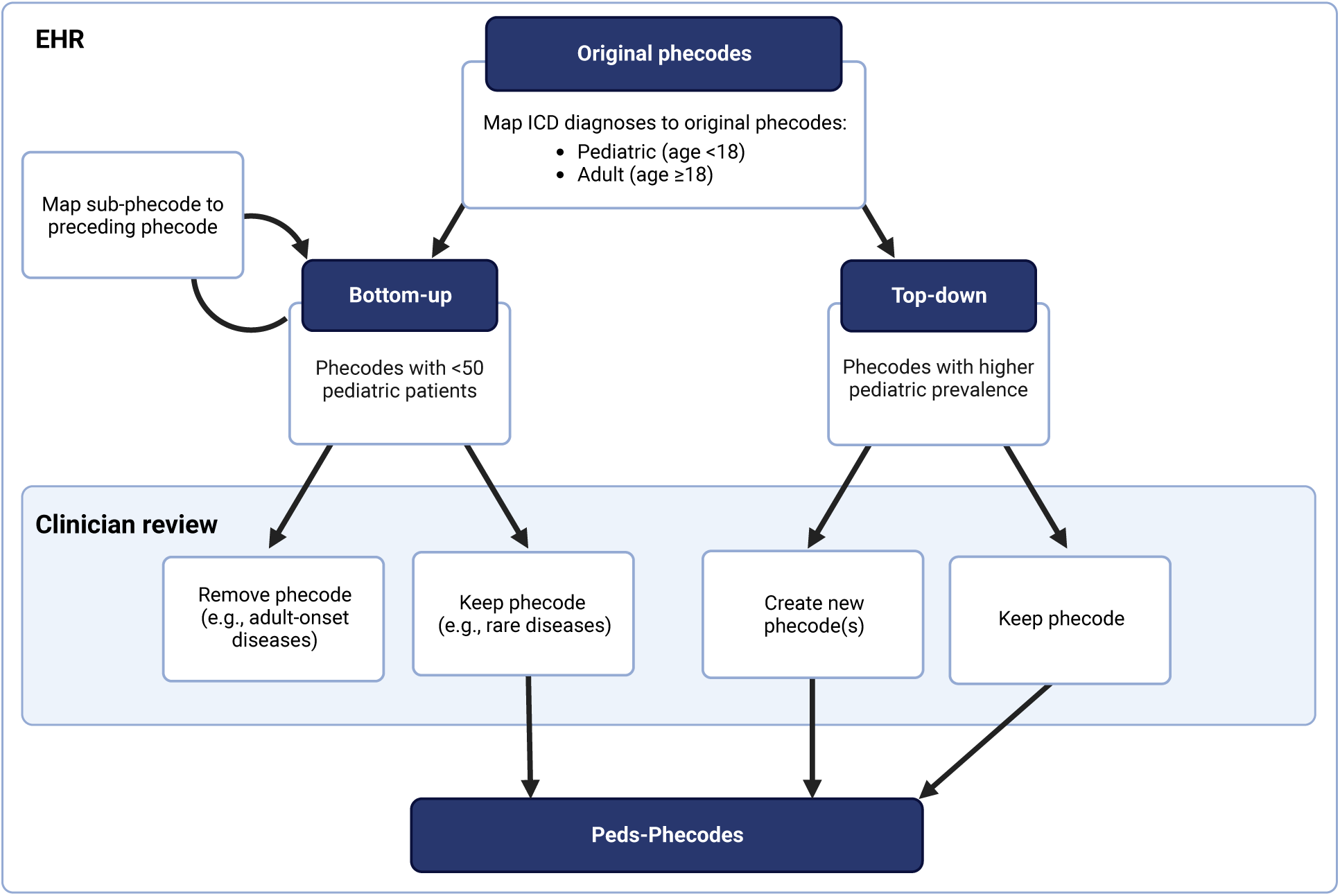
Workflow for Peds-Phecodes development. We used a hybrid data- and knowledge-driven approach combining patient diagnosis counts from VUMC with clinician-led manual review to prune out phenotypes with little pediatric relevance (left) and create new phecodes for important pediatric phenotypes (right).

From bottom-up, we pruned the phecodes without sufficient pediatric patient representation to identify potential phecodes for removal. We iteratively remapped each sub-phecode with <50 pediatric patients (from a study population of >1 million pediatric patients) to the phecode directly above it in the phecode hierarchy, starting at the most granular level of the hierarchy (level 2 sub-phecodes) and working up to the root phecode. In this way, we collapsed uncommon sub-phecodes while preserving the underlying hierarchical structure of the phecodes. For example, sub-phecodes 153.2 “Colon cancer” and 153.3 “Malignant neoplasm of rectum, rectosigmoid junction, and anus” were consolidated into their root phecode 153 “Colorectal cancer”. Two pediatricians (SLV and PJK) reviewed the remaining low count phecodes to identify those that represented diseases not applicable to the pediatric population (e.g., phecode 453 “Chronic venous hypertension” and phecode 796 “Elevated prostate specific antigen”), which were then removed. Low pediatric count phecodes representing rare diseases (e.g., phecode 209 “Neuroendocrine tumors”) were preserved.

From top-down, we focused on the phecodes with significantly higher prevalence in our pediatric cohort. For each phecode with higher pediatric prevalence, the two pediatricians reviewed the distribution of pediatric and adult patient diagnoses mapped to the phecode and provided recommendations for new phecodes reflecting diseases of particular importance to the pediatric population (Supplementary Figure 2). To provide a clearer picture of the intra-phecode ICD distributions, ICD-10-CM diagnoses were converted to ICD-9-CM using the General Equivalence Mappings (GEMS) provided by the Centers for Medicare & Medicaid Services.[14] Both the ICD-9-CM codes and matching ICD-10-CM codes were remapped to the newly created phecodes. We also mapped several ICD-10-CM codes without ICD-9-CM analogs (e.g., ICD-10-CM codes related to COVID-19) to new phecodes.

### PheWAS analyses

To validate the Peds-Phecodes, we conducted PheWAS analyses using Peds-Phecodes and the current phecodes independently and compared their ability to replicate known genotype-phenotype associations from previous pediatric studies. We queried the NHGRI-EBI GWAS Catalog[15] to find genetic variants associated with pediatric phenotypes for our PheWAS replication studies. We identified 687 SNPs in the GWAS Catalog that could be investigated using VUMC’s DNA biobank. These SNPs were associated with eight different pediatric phenotypes: congenital heart disease, pyloric stenosis, Hirschsprung disease, hypospadias, café-au-lait spots (observed in neurofibromatosis type I), pediatric eosinophilic esophagitis, childhood-onset asthma, and juvenile idiopathic arthritis. We performed PheWAS analyses for all 687 SNPs using binary logistic regression, adjusting for sex and race. We only used diagnoses made during the pediatric age window (i.e., ICD-9-CM and ICD-10-CM codes recorded at <18 years of age) in creating the phenotypes for PheWAS. We required a minimum of two ICD codes (both recorded at <18 years of age) for a patient to be counted as a case.

As in previous studies, replication was defined as the detection of signals related to the phenotype of interest with concordant effect directionality and p<0.05.[6] We evaluated replication using both an exact and approximate phenotype match. In the exact phenotype match, we focused only on the PheWAS signal for the singular phecode best representing the phenotype of the genetic association from the GWAS Catalog (e.g., Peds-Phecode 747.1113 “Transposition of great vessels” and phecode 747.13 “Congenital anomalies of great vessels” for replication of the association between rs150246290 and transposition of the great arteries[16]). In the approximate match, we broadened our replication phenotype, examining the signals for all sub-phecodes stemming from the root phecode best matching the phenotype of the known association. For example, the approximated replication phenotype for the association between rs150246290 and transposition of the great arteries was represented by root phecode 747 “Cardiac and circulatory congenital anomalies” and all its sub-phecodes (N = 57 sub-phecodes in Peds-Phecodes, N = 5 in the current phecodes).

Beyond replication, we also evaluated the ability of the Peds-Phecodes to detect novel genetic associations compared to the existing phecodes. We compared the overlapping and unique signals generated in each PheWAS and examined the significant signals detected using Peds-Phecodes but not phecodes (p<Bonferroni-corrected p).

We developed the PedsPheWAS R package to facilitate PheWAS analysis using the Peds-Phecodes (https://github.com/The-Wei-Lab/PedsPheWAS).

## RESULTS

### Differences in disease prevalence in the pediatric and adult populations

We observed marked differences in the distribution of disease between the pediatric and adult populations. Phecode 465 “Acute upper respiratory infections of multiple or unspecified sites” represented the most common overall pediatric disease phenotype, with a prevalence of 26.4% in the pediatric population compared to a prevalence of 16.9% in the adult population (p<0.001), followed by phecode 656 “Other perinatal conditions of fetus or newborn” (pediatric prevalence = 23.7%, adult prevalence = 1.7%, p<0.001). In contrast, the most common overall disease phenotype among the adult population was phecode 401 “Hypertension” (adult prevalence = 31.8%, pediatric prevalence = 1.8%, p<0.001).

Consistent with our hypothesis that pediatric and adult patients are differentially affected by disease, we found that nearly all phecodes (1,783/1,866, or 95.6%) differed significantly in prevalence in the pediatric and adult populations. We identified 317 phecodes with higher prevalence among pediatric patients at VUMC compared to adult patients and 1,466 phecodes with lower prevalence in pediatric patients compared to adults (Bonferroni-corrected p = 0.05/1,866 = 2.7 x 10^-5^) (Supplementary Tables 1-2). The phecodes with significantly higher pediatric prevalence included congenital anomalies (N = 51), disorders affecting the sense organs (N = 30), diseases affecting the digestive system (N = 26), and injuries and poisonings (N = 26). Among the phecodes with significantly higher prevalence in the adult population were diseases of the circulatory system (N = 164), genitourinary conditions (N = 157), and endocrine/metabolic conditions (N = 136). Figure 2 shows the top five level 2 sub-phecodes with significantly higher prevalence in the pediatric population, ordered by descending prevalence, along with the corresponding sub-phecodes with significantly higher prevalence in the adult population.

**Figure 2.**
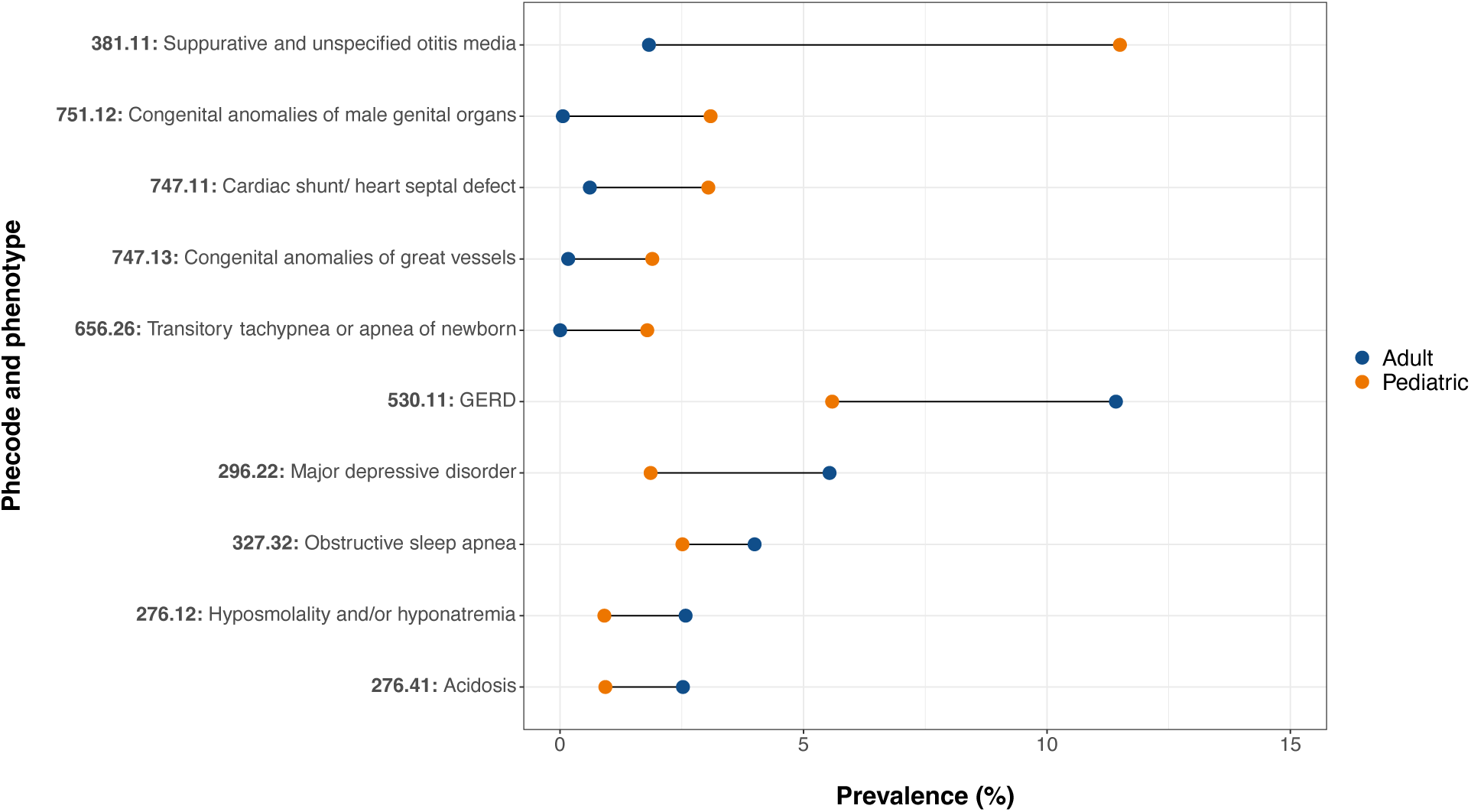
Comparison of phecode prevalence between the pediatric and adult populations at VUMC. Patient diagnoses at <18 years of age were mapped to the most recent phecodes version. The top five phecodes with significantly higher pediatric prevalence are displayed in the top half the dumbbell plot; the top five phecodes with significantly higher prevalence among adults are shown in the bottom half. Because of the multiple levels of phecode granularity available, direct comparisons can only be made between phecodes existing at the same level of the phecode hierarchy. Here, we show results for the level 2 (i.e., most granular) sub-phecodes in the current phecode hierarchy.

### Pediatric phecodes (Peds-Phecodes)

The final Peds-Phecodes set consists of 2,051 phecodes, compared with 1,866 phecodes in the most recent phecodes iteration. The 2,051 Peds-Phecodes are subdivided into 586 root phecodes with five possible levels of sub-phecodes (N = 1,016 level 1 sub-phecodes, N = 322 level 2, N = 106 level 3, N = 18 level 4, and N = 3 level 5), in contrast to the 1,866 phecodes, which contain 585 root phecodes with two possible levels of sub-phecodes (N = 1,012 level 1, N = 269 level 2). These 2,051 phecodes provide a mapping for 15,533 ICD-9-CM codes and 82,949 ICD-10-CM codes. As the pediatric phecodes were developed using the existing phecodes as a starting point, there is still much overlap between the two, with 1,693 shared phecodes (Figure 3). In developing the Peds-Phecodes, 173 phecodes were removed from the starting phecode set and 358 new phecodes were created. The majority of the new phecodes created (351/358) were sub-phecodes of existing root phecodes, resulting in an extension of the phecode hierarchy to a maximum of six levels rather than three.

**Figure 3.**
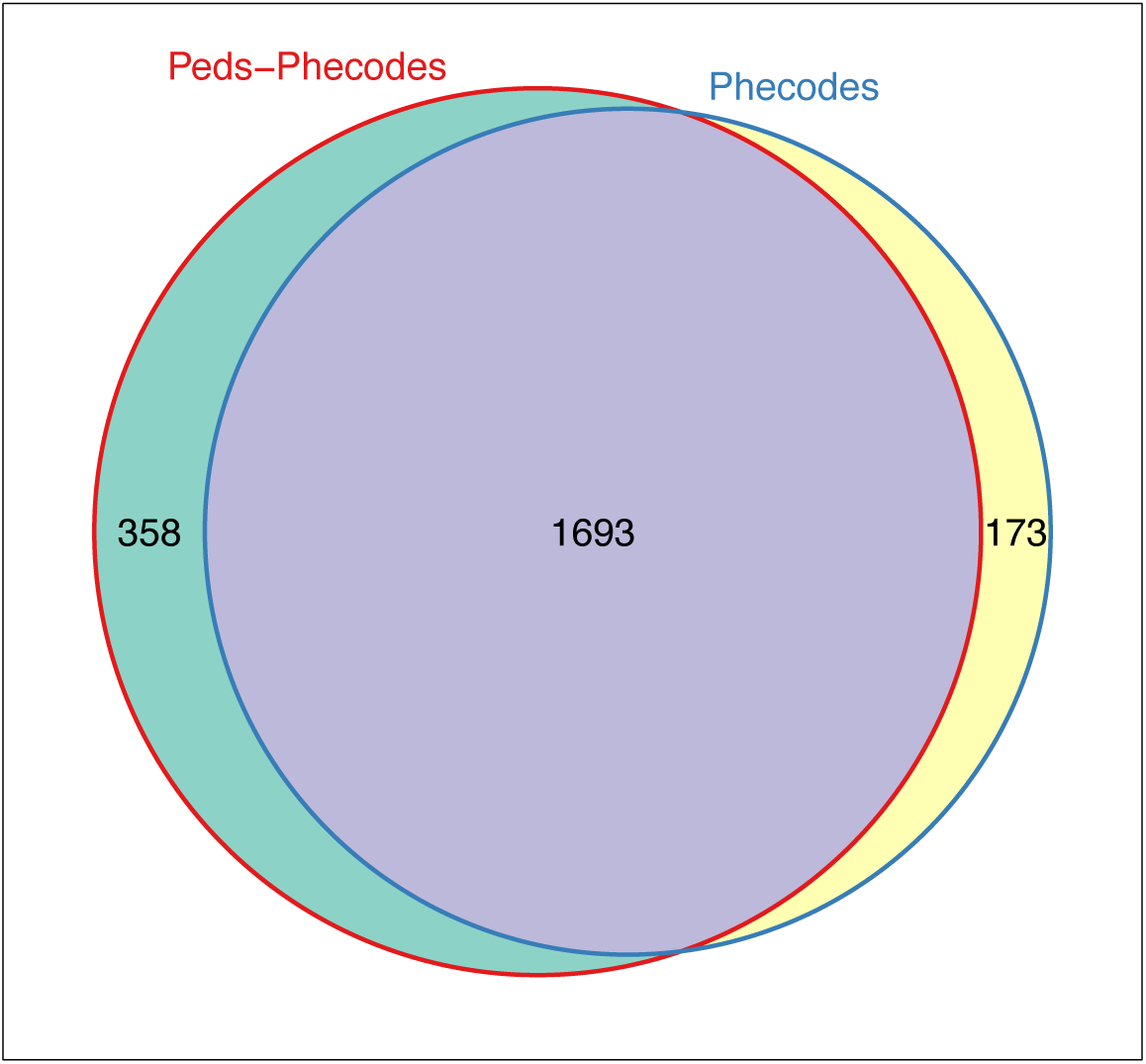
Comparison of Peds-Phecodes and phecodes. Weighted Venn diagram showing the overlap between Peds-Phecodes and phecodes.

Furthermore, the Peds-Phecodes were recategorized into 19 disease categories: circulatory system, dermatologic, endocrine/metabolic, digestive, genitourinary, hematologic, musculoskeletal, neurological, renal/kidney, respiratory, sense organs, congenital anomalies and genetic disorders, infections, immunologic and inflammatory disorders, neoplasms, mental disorders, pregnancy complications, symptoms, and injuries and poisonings. The renal/kidney and immunologic and inflammatory disorders categories were newly created for the Peds-Phecodes. We sought to group the Peds-Phecodes into disease categories based on the affected anatomical location – for instance, Peds-Phecode 747 “Cardiac and circulatory congenital anomalies” was placed into the circulatory system disease category, although phecode 747 belongs to the congenital anomalies category in phecodes version 1.2.

### Replication of known genotype-phenotype associations

Considering only the signal for the singular phecode best matching the phenotype of interest (i.e., exact phenotype match), use of the Peds-Phecodes replicated 88/687 (12.8%) of the SNP-phenotype associations extracted from the GWAS Catalog with p<0.05, compared to 80/687 (11.6%) replicated using the phecodes. Given this relatively low replication rate for both phecode sets in the context of limited pediatric GWAS availability and the smaller pediatric sample sizes for PheWAS, we expanded our definition of the replication phenotype. Rather than using a single phecode as the replication phenotype, we defined the replication phenotype using the root phecode best matching the phenotype of interest as well as all its sub-phecodes (i.e., approximate phenotype match).

Using the approximate phenotype match, we successfully replicated 248/687 (36.1%) associations at p<0.05 using the Peds-Phecodes, a significant increase from the 192/687 (27.9%) replicated using the phecodes (p<0.001 using McNemar’s exact test) (Supplementary Tables 3-4). Additionally, use of the Peds-Phecodes generally yielded smaller p-values for known genotype-phenotype associations compared to the phecodes, despite similar statistical power based on the number of identified cases (Figure 4).

**Figure 4.**
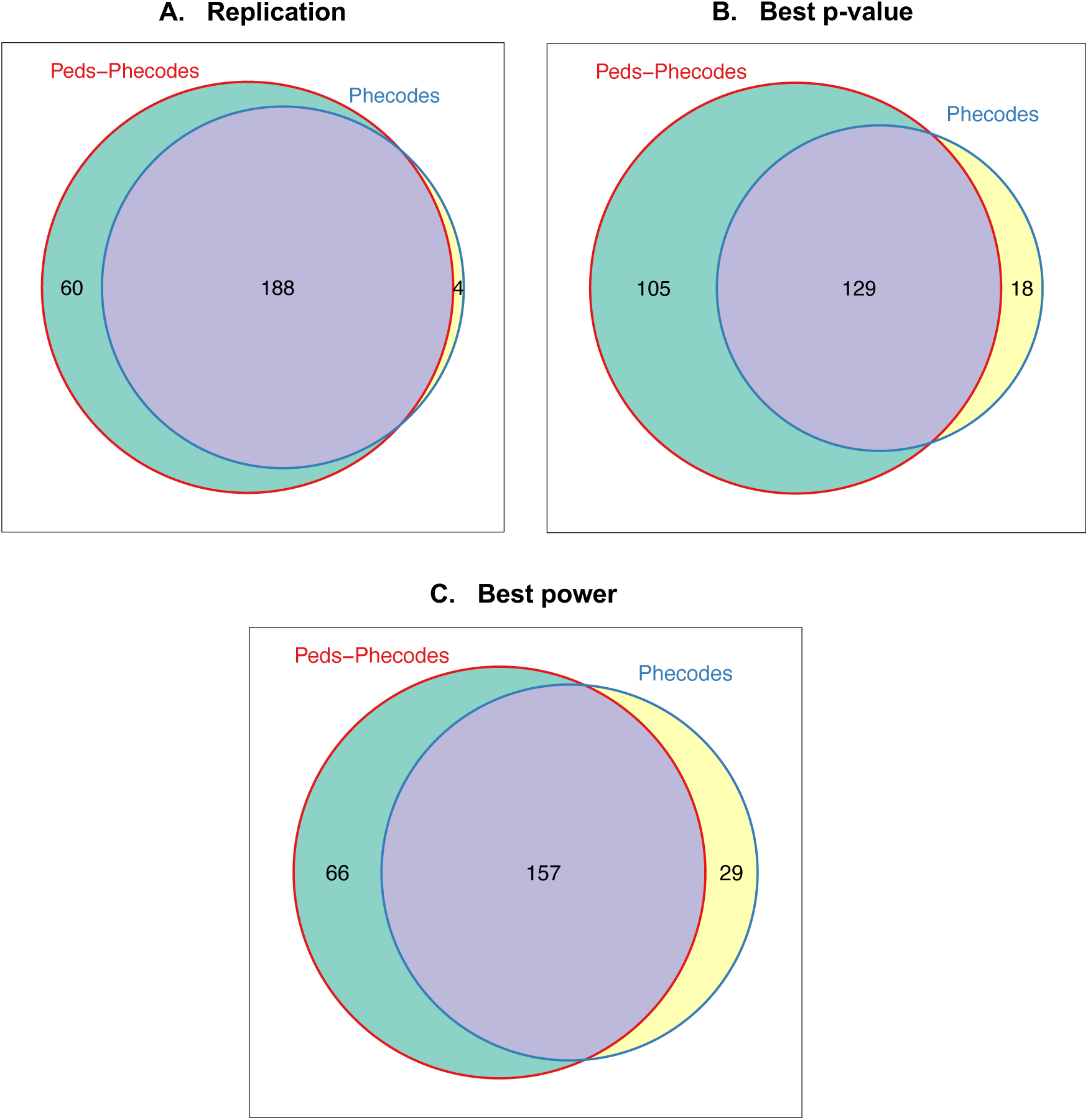
Performance of Peds-Phecodes and phecodes in PheWAS replication studies. Weighted Venn diagrams comparing Peds-Phecodes and phecodes in terms of their ability to replicate known genotype-phenotype associations (panel A), the replication p-values (panel B), and statistical power (panel C).

Using Peds-Phecodes, we were able to replicate several genotype-phenotype associations that we were unable to replicate using the phecodes. In Figure 5, we show the results of our PheWAS for SNP rs2505998, a genetic variant in the *RET* gene, which has previously been shown to be associated with Hirschsprung’s disease, a congenital defect of the intestine that most commonly affects the colon and rectum.[17] The PheWAS performed using Peds-Phecodes captured the expected association between phecode 750.214 “Hirschsprung’s disease and other congenital functional disorders of colon” (OR = 2.23, p = 6.02 x 10^-4^); however, the PheWAS using the phecodes was not able to exactly replicate the association (OR = 1.13, p = 0.22);. Our PheWAS analyses also reflected the higher granularity of phenotypes available through the Peds-Phecodes. In the PheWAS for rs138741144, an *ASIC2* variant previously associated with cardiac septal defects,[16] using Peds-Phecodes revealed significant associations with phecode 747.113 “Common ventricle”, phecode 747.128 “Atresia and stenosis of aorta”, phecode 747.13 “Congenital anomalies of great vessels”, and phecode 747.133 “Patent ductus arteriosus”. In contrast, the PheWAS using the phecodes produced a much less granular signal, with only one significant association related to congenital heart disease: phecode 747.13 “Congenital anomalies of great vessels” (Figure 6).

**Figure 5.**
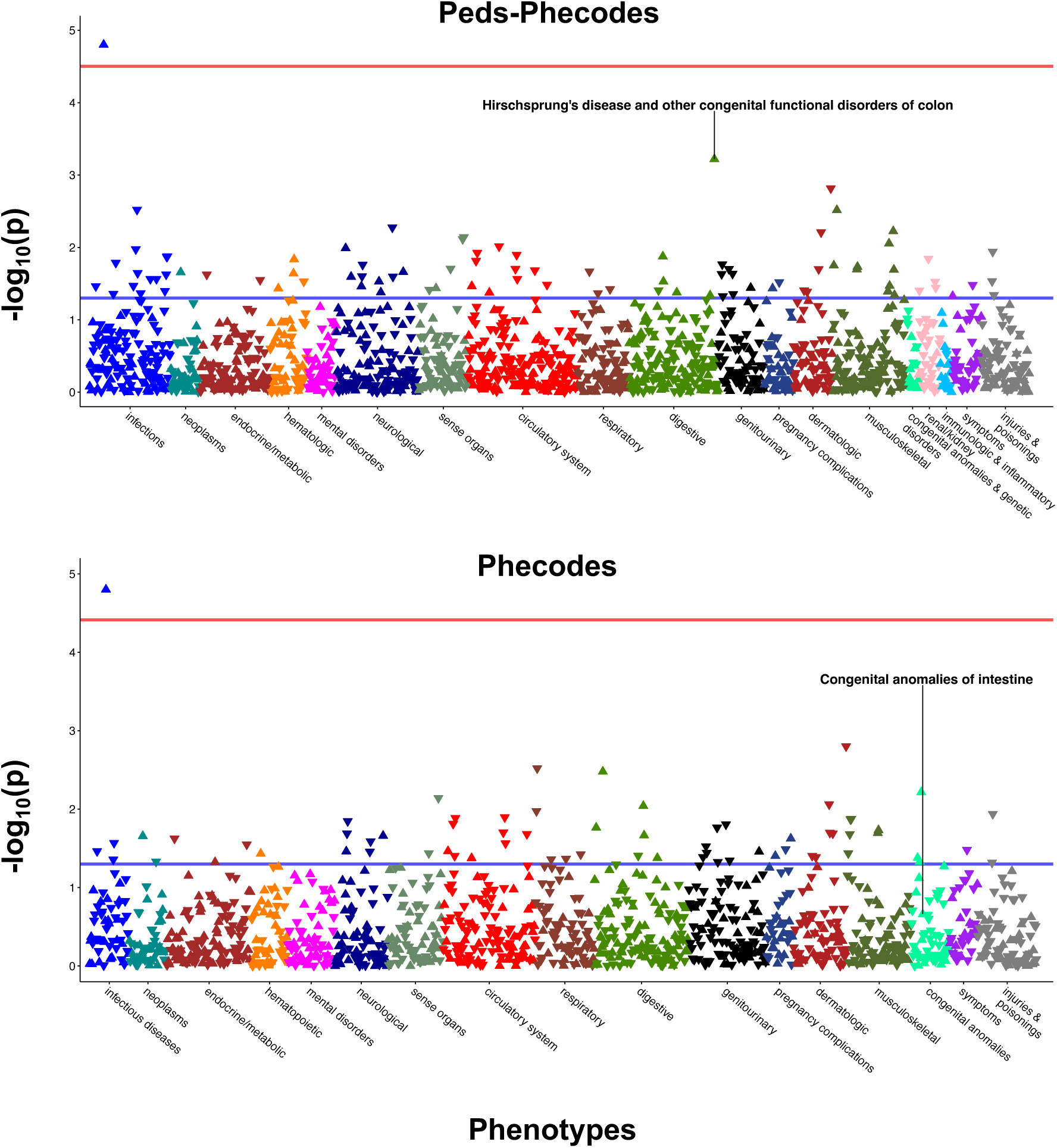
Comparative PheWAS analyses for rs2505998, which has previously been associated with Hirschsprung’s disease. PheWAS was performed using Peds-Phecodes (top) and phecodes (bottom). The horizontal blue line indicates p = 0.05; the red line indicates the Bonferroni-corrected p-value threshold. In both analyses, the most significant PheWAS association for rs2505998 was observed for phecode 070.2 “Viral hepatitis B”.

**Figure 6.**
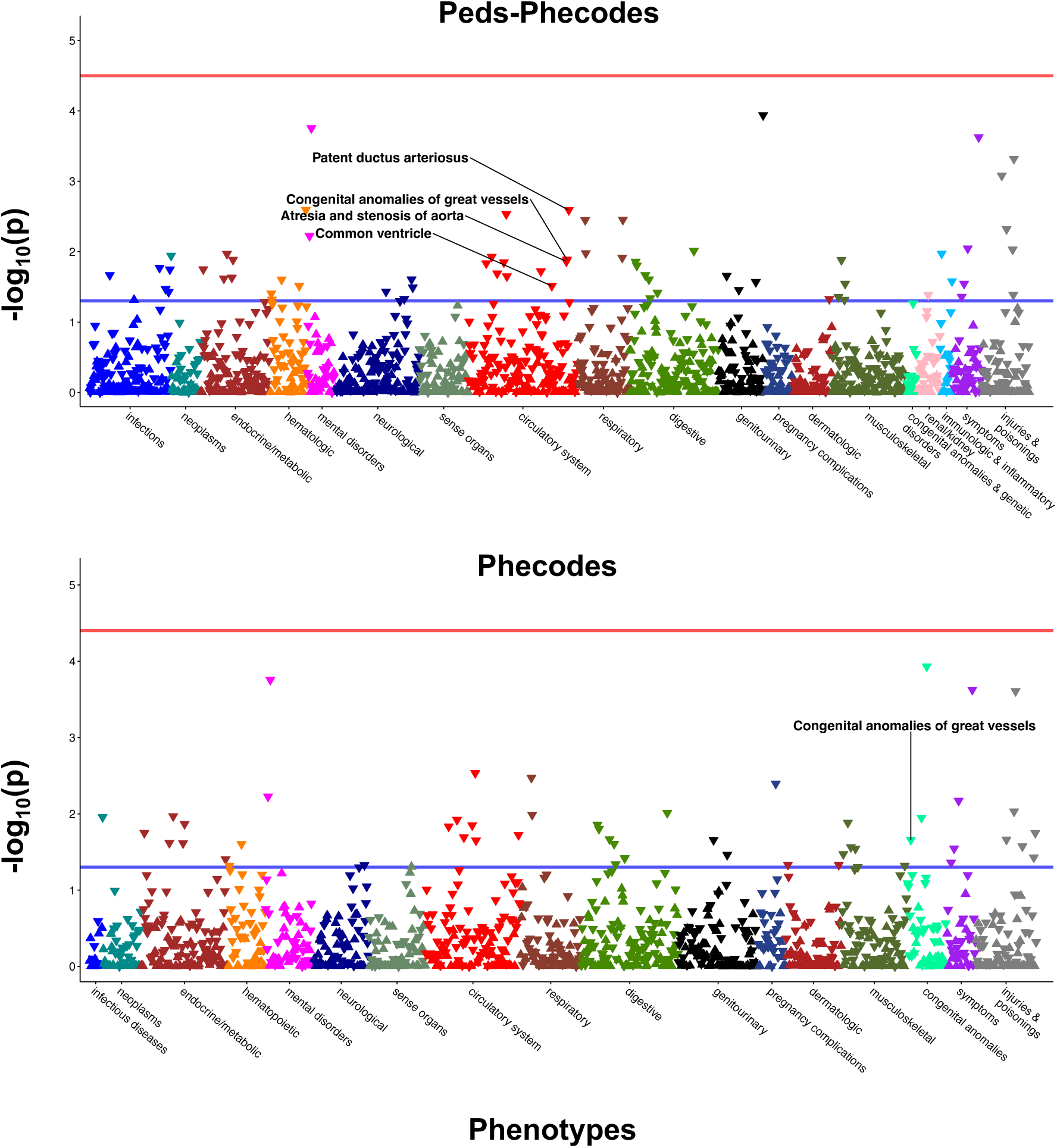
Comparative PheWAS analyses for rs138741144, which has previously been associated with heart septal defects. PheWAS was performed using Peds-Phecodes (top) and phecodes (bottom). The horizontal blue line indicates p = 0.05. No signals cross the Bonferroni-corrected p-value threshold (red line).

We also examined the overlap between the top signals produced in PheWAS using Peds-Phecodes compared to phecodes. For 99.7% (685/687) of SNP-phenotype pairs, the PheWAS analysis using Peds-Phecodes resulted in a larger number of signals with p<0.05 compared to the PheWAS using the phecodes. We found a significant difference between the number of significant signals in the Peds-Phecodes PheWAS compared to the phecode PheWAS using a paired t-test (p<0.001). The number of overlapping signals with p<0.05 ranged from 2.3% (16/687) to 29.5% (203/687). For each genotype-phenotype association, the PheWAS using Peds-Phecodes revealed several signals (minimum N = 3, maximum N = 75) that were not detected using the phecodes, representing potentially novel SNP-phenotype associations.

To further explore the novel associations identified using the Peds-Phecodes, we examined the unique signals detected in the Peds-Phecodes PheWAS analyses applying a Bonferroni-corrected significance threshold rather than a replication p-value threshold. We found that the Peds-Phecodes identified novel associations for 147/687 (21.4%) of the SNPs tested in PheWAS, with a total of 348 novel associations detected (maximum N = 16 for the *TLR1* variant rs5743618) (Supplementary Table 5). Interestingly, these new associations were not limited to new phecodes created when developing the Peds-Phecodes from the existing phecodes – we identified new genetic associations for 55 distinct phenotypes, 27 of which were phecodes shared by Peds-Phecodes and the phecodes. For example, we identified 97 new genetic associations for phecode 465.3 “Acute upper respiratory infection” (a new phecode) and 43 new associations for phecode 079 “Viral infection” (a shared phecode).

## DISCUSSION

The Peds-Phecodes represent a first attempt to develop a high-throughput EHR phenotyping tool specifically designed for pediatric research, with the aim of facilitating large-scale pediatric-specific phenome-wide and genome-wide studies by streamlining and standardizing the phenotyping process in a manner that reflects the unique pediatric spectrum of disease.

EHRs provide an archive of longitudinal clinical data with vast potential for research use. However, defining phenotypes efficiently and accurately for research within the EHR remains a challenging and nuanced process. Phecodes emerged from an effort to characterize the phenome by creating standardized groupings of ICD codes to represent clinically meaningful phenotypes, making it possible to perform PheWAS analyses quickly and effectively using EHR data. However, phecodes were developed using population-based diagnoses predominantly affecting adult patients, and thus are best suited for cohorts primarily composed of adult patients.

When we mapped patient diagnoses in VUMC’s EHR to the current phecodes version, we observed distinct differences in the diagnoses recorded while patients were <18 years of age (i.e., pediatric patients) and those recorded after patients were ≥18 years of age. As shown in Figure 2, phecodes related to infections (e.g., otitis media) and congenital anomalies had higher prevalence in the pediatric population, whereas chronic diseases (e.g., gastroesophageal reflux disease, obstructive sleep apnea, and major depressive disorder) were more common among adults. These differences reflect the unique distributions of disease among pediatric and adult populations and highlight the need for modified phecodes for pediatric-specific PheWAS. Other studies have reported similar differences in patterns of disease among children and adults,[18,19] providing further motivation for specialized pediatric phecodes.

We developed the pediatric phecodes using a hybrid data- and knowledge-driven approach – we leveraged diagnosis code counts to help us approximate the importance (or lack thereof) of each existing phecode in the pediatric population. Diagnosis codes provided a consistent and replicable way of performing the initial assessment and significantly reduced manual review efforts. Clinical domain knowledge then fine-tuned the detected signals and ascertained their practical importance. With this insight, we modified the existing phecodes to capture diseases primarily affecting pediatric patients with increased granularity and exclude diseases not relevant to pediatric patients, capturing clinically meaningful pediatric phenotypes. The results of our PheWAS analyses reflect the ability of Peds-Phecodes to better capture pediatric conditions. Using Peds-Phecodes, we replicated more of the genotype-phenotype associations extracted from the GWAS Catalog compared to the phecodes. We also found that using the Peds-Phecodes tended to produce smaller p-values in our PheWAS replication studies.

Peds-Phecodes substantially increased the granularity with which certain phenotypes could be characterized. In developing Peds-Phecodes, we did not remove the underlying hierarchical structure of phecodes; rather, we further extended the hierarchy vertically to accommodate a maximum of five levels of added granularity rather than two. We also extended the phecode hierarchy horizontally, incorporating alphanumeric codes to allow a phecode to directly branch into more than nine possible sub-phecodes (e.g., phecode 758.1A “Gonadal dysgenesis”). The results of our comparative PheWAS analyses highlight the advantages of the added granularity built into the Peds-Phecodes. When attempting to find an exact phenotype match for the replication studies, phecodes tended to represent phenotypes that were much broader than the target phenotype. For instance, phecodes 751.12 “Congenital anomalies of male genital organs” and 750.21 “Congenital anomalies of intestine” were the best matches for hypospadias and Hirschsprung’s disease, respectively. However, the Peds-Phecodes directly represented these phenotypes (phecodes 751.124 “Hypospadias” and 750.214 “Hirschsprung’s disease and other congenital functional disorders of colon”). Using an approximate phenotype match, we were able to replicate more known genotype-phenotype associations using the Peds-Phecodes largely because of their increased granularity.

Differences in the significant signals obtained in PheWAS using Peds-Phecodes and phecodes also revealed the potential of the Peds-Phecodes to detect novel associations. Although we found at least sixteen overlapping phecodes with p<0.05 in each PheWAS comparison, we also found at least three signals with p<0.05 unique to each Peds-Phecodes PheWAS for each SNP. Furthermore, we identified a total of 348 novel SNP-phenotype associations with Bonferroni significance using the Peds-Phecodes. These signals warrant further investigation, and we hope to further explore the novel associations identified using the Peds-Phecodes in large-scale pediatric PheWAS analyses in future studies.

This study has several limitations. While we created over 350 new phecodes in developing the Peds-Phecodes, we faced difficulties in finding SNPs associated with pediatric phenotypes for PheWAS validation, particularly in the realm of infectious diseases. This may in part have been due to our reliance on the GWAS Catalog to identify genetic associations; use of other repositories of genotype-phenotype data (e.g., Online Mendelian Inheritance in Man), particularly those focusing specifically on rare diseases, may have yielded other genetic variants for testing, and should be explored in subsequent studies. Still, pediatric patients are comparatively understudied in GWAS and the pediatric-specific studies available tend to focus on a relatively small number of conditions, limiting the breadth of the replication PheWAS analyses that we were able to perform. We extracted 687 SNPs associated with eight pediatric phenotypes from the GWAS Catalog. Four of these eight pediatric phenotypes were congenital anomalies, two were pediatric autoimmune diseases, one was a common inflammatory disease (asthma), and one was a rare disease (neurofibromatosis type I), providing some level of variety but nothing close to comprehensive coverage of the pediatric phenome. Our replication analyses were also limited by smaller sample sizes resulting in lower statistical power, as there are less pediatric data contained within EHRs. However, pediatric EHR data will accumulate over time, paving the way for more high-powered PheWAS analyses in pediatric cohorts in the future. Finally, we acknowledge that the ICD code to phecode mapping will likely always have room for improvement. Knowledge assembly is an iterative process – as researchers apply the pediatric phecodes in their work, we hope to incorporate their feedback and continue to update the mapping. We will also continue to extend the pediatric phecode map to include newly added ICD-10-CM codes.

The Peds-Phecodes can be browsed and downloaded at: https://wei-lab.app.vumc.org/phecode-data/pediatric_phecodes

## CONCLUSION

There is an urgent need for high-throughput EHR phenotyping tools specialized for use in pediatric populations. Building on the existing phecodes, we developed new pediatric phecodes (Peds-Phecodes) using a hybrid approach integrating diagnosis data and clinical domain knowledge. Using the Peds-Phecodes to conduct PheWAS analyses, we were able to replicate significantly more known genotype-phenotype associations for pediatric conditions compared to using the phecodes, as well as identify potentially novel genotype-phenotype associations. We have made a PedsPheWAS R package to perform PheWAS using the Peds-Phecodes.

## Data Availability

All data produced in the present study are available upon reasonable request to the authors and subsequent institutional approval.

https://wei-lab.app.vumc.org/phecode-data/pediatric_phecodes

https://wei-lab.app.vumc.org/phecode-data/phecodes

## SUPPLEMENTARY MATERIAL

**Supplementary Figure 1.**
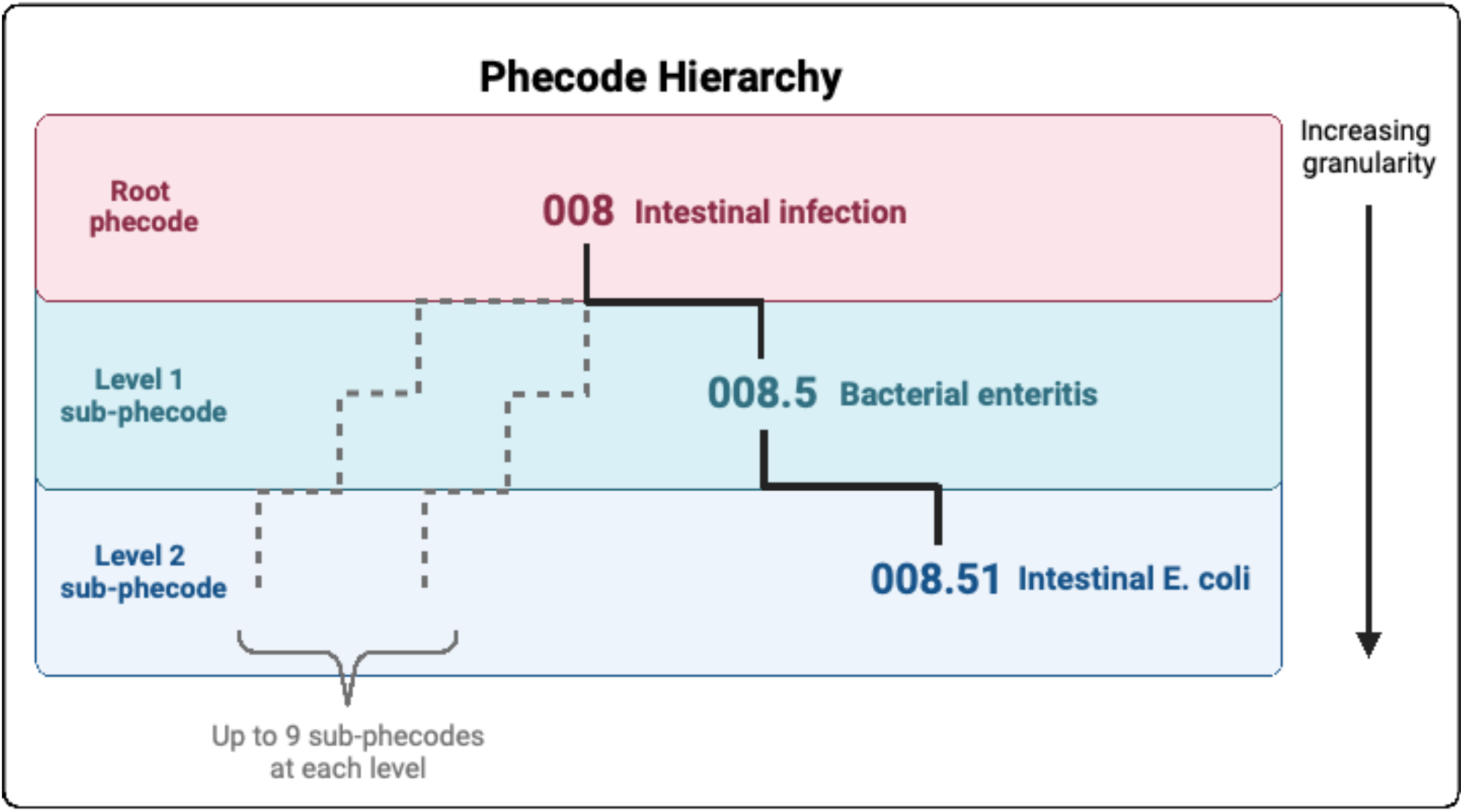
Representative example of the phecode hierarchy. Phenotypic granularity progressively increases moving down the hierarchy from the root phecode (least granular) to level 2 sub-phecodes (most granular).

**Supplementary Figure 2.**
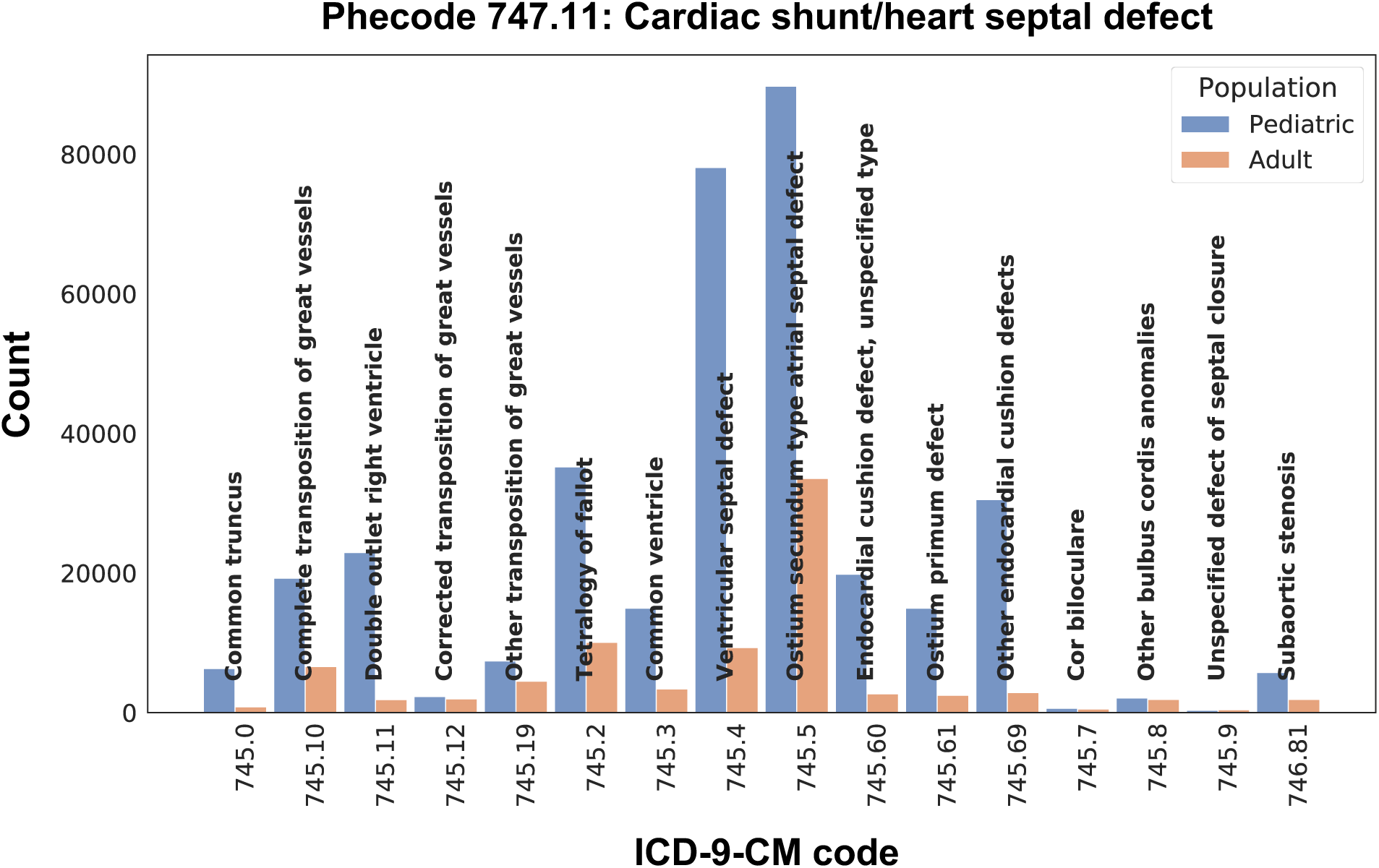
Representative example of new phecode creation. Based on patient counts for ICD codes originally mapped to phecode 747.11 “Cardiac shunt/heart septal defect”, new sub-phecodes were created in the pediatric version (e.g., Peds-Phecode 747.112 “Tetralogy of Fallot”).

## ACKNOWLEDGEMENTS

The authors wish to acknowledge the MPRINT Steering Committee for its support.

